# RF shimming strategy for an open 60-channel RF transmit 7T head coil for routine use on the single transmit mode

**DOI:** 10.1101/2024.11.13.24316383

**Authors:** Andrea N Sajewski, Tales Santini, Anthony DeFranco, Jacob Berardinelli, Hecheng Jin, Jinghang Li, Cong Chu, Jeremy J Berardo, Tamer S Ibrahim

## Abstract

**Purpose:** To develop an RF shimming approach for operating the 2^nd^ Generation Tic Tac Toe RF Coil System (60 transmit channels integrated with 32-channel receive insert) for routine use in 7T neuro MRI on the single transmit mode.

**Methods:** RF simulations were performed and used to develop non-subject-specific RF shim cases over three anatomically detailed head models: adult male, adult female, and child female. Multi-ROI shimming strategies were developed and implemented. B_1_^+^ maps and in-vivo images were acquired on the single transmit mode of a 7T scanner using the RF shim cases derived from the computer simulations.

**Results:** The availability of 60 transmit channels enables more control over RF power efficiency, SAR efficiency, and B_1_^+^ homogeneity using RF shimming. On the single transmit mode, the 2^nd^ Generation Tic Tac Toe RF Coil System consistently provides homogeneous B_1_^+^ field distribution with extended coverage into the temporal lobes, cerebellum, reaching all the way to C5-C6. Safe levels of SAR are also achieved.

**Conclusion:** By using a non-subject specific RF shimming approach derived from computer simulations, the 2^nd^ Generation Tic Tac Toe RF Coil System allows for robust, routine neuroimaging (> 1750 in-vivo scanning sessions over the past 28 months) at 7T in single transmit mode.

## Introduction

Ultra-high field (UHF) human MRI, designated as having a primary magnetic field (B_10_) strength of 7T or higher, has shown clinical utility in neuro and musculoskeletal imaging^1,2^, offering improvements in signal-to-noise ratio^3,4^, contrast enhancement, spatial and spectral resolution, and BOLD sensitivity^5,6^. With FDA clearance for multiple vendors in recent years^7,8^, 7T sites have been increasing for clinical and research use.

Despite its advantages, UHF MRI still poses technical challenges because of the increased operational frequency and thus reduced wavelength (∼13 cm for in-vivo proton neuroimaging at 7T). This shortened wavelength leads to spatial inhomogeneity of the radiofrequency (RF) fields^3,9^: inhomogeneity in the transmitted magnetic field (B_1_^1+^) may lead to signal dropout in the MR images, while electric field inhomogeneity may lead to increased localized tissue heating quantified by specific absorption rate (SAR). Additionally, UHF MRI systems lack a built-in body transmit coil, requiring custom or commercial transmit coils for neuroimaging. The scarcity of commercial systems and the challenge of achieving consistent whole-brain homogenous excitation has driven research groups to develop custom transmit coils^10,11^.

One approach to address spatial inhomogeneity is to utilize a multichannel transmit system, in which each transmit channel can be driven independently and the resultant RF fields can be combined with superposition^12^. RF shimming, the process of optimizing the amplitudes and phases of a multichannel transmit system, is used to help mitigate the inhomogeneity of the electric and magnetic fields^13,14^. This can be done on a per-subject basis using a parallel transmission (pTx) system, which has been able to provide more homogeneous B_1_^+^ fields^15^, but may pose higher risk due to challenges in calculating SAR per subject, has longer setup times, and was only recently FDA cleared for clinical use^16^. For sites with existing 7T systems, the single-transmit (sTx) mode of the scanner is often used, requiring non-subject-specific RF transmit systems. However, sTx coils based on the traditional quadrature birdcage design, which are widely used, usually struggle to provide sufficient B_1_^+^ intensity to the entire brain, especially in the temporal lobes and cerebellum^17,18^. The use of dielectric pads could potentially improve the signal in these lower brain regions^17,18^; but this comes at the cost of SAR changes and/or inducing field inhomogeneities in other regions^18,19^. Furthermore, to improve B_1_^+^ efficiency for UHF MRI, many coil designs are fully covered in front of the face and visual field, not only creating challenges with patient comfort and usability, but also limiting the viewing of visual stimuli as is often required for functional MRI experiments.

The 1^st^ generation Tic-Tac-Toe (TTT) design, established previously^20-22^, demonstrated the ability to provide a homogeneous and load-insensitive B_1_^+^ field for neuro MRI at 7T in sTx mode with 16 combined transmit channels^23-25^. In this work, we examine RF shimming on the next-generation TTT transmit coil^26^. By increasing the number of transmit channels, the degrees of freedom in RF shimming increases, allowing for improvements in homogeneity and extended coverage. We analyzed non-subject-specific RF shimming strategies for this multi-channel transmit array for use with sTx systems. We performed finite difference time domain (FDTD) simulations and numerical optimizations over three anatomically detailed head models to develop three different RF shim cases, each of which was experimentally implemented and tested in-vivo on 7T. By shimming over the three head models simultaneously, we were able to achieve a robust coil operation in sTx mode for routine neuroimaging at 7T.

## Methods

### Tic-Tac-Toe Design

The TTT panel consists of eight transmission lines connected to each other, with half used as RF inputs and half for frequency tuning^21-25^. Solid copper rods are inserted into each leg of the dielectric TTT structure to create the coaxial transmission line and can be adjusted for tuning and matching; capacitors are used for further tuning of the coil (Figure 1c, d). In this design, each TTT panel is 98 mm x 98 mm x 23 mm for the inner copper shield. The distance between the strut and the shield was optimized for homogeneity and SAR reduction^27^. A total of 15 TTT panels are arranged in a two-row octagonal array around the head (Figure 1a, b), providing a total of 60 transmit channels. The coil is open in front of the face from eye level to promote patient comfort and to allow easy access to fMRI projection. The shielding extends 144mm beyond the top row of the coil to accommodate the preamplifier boards for the receive insert and is slanted to increase the field of view for fMRI stimulus projection.

**Figure 1:**
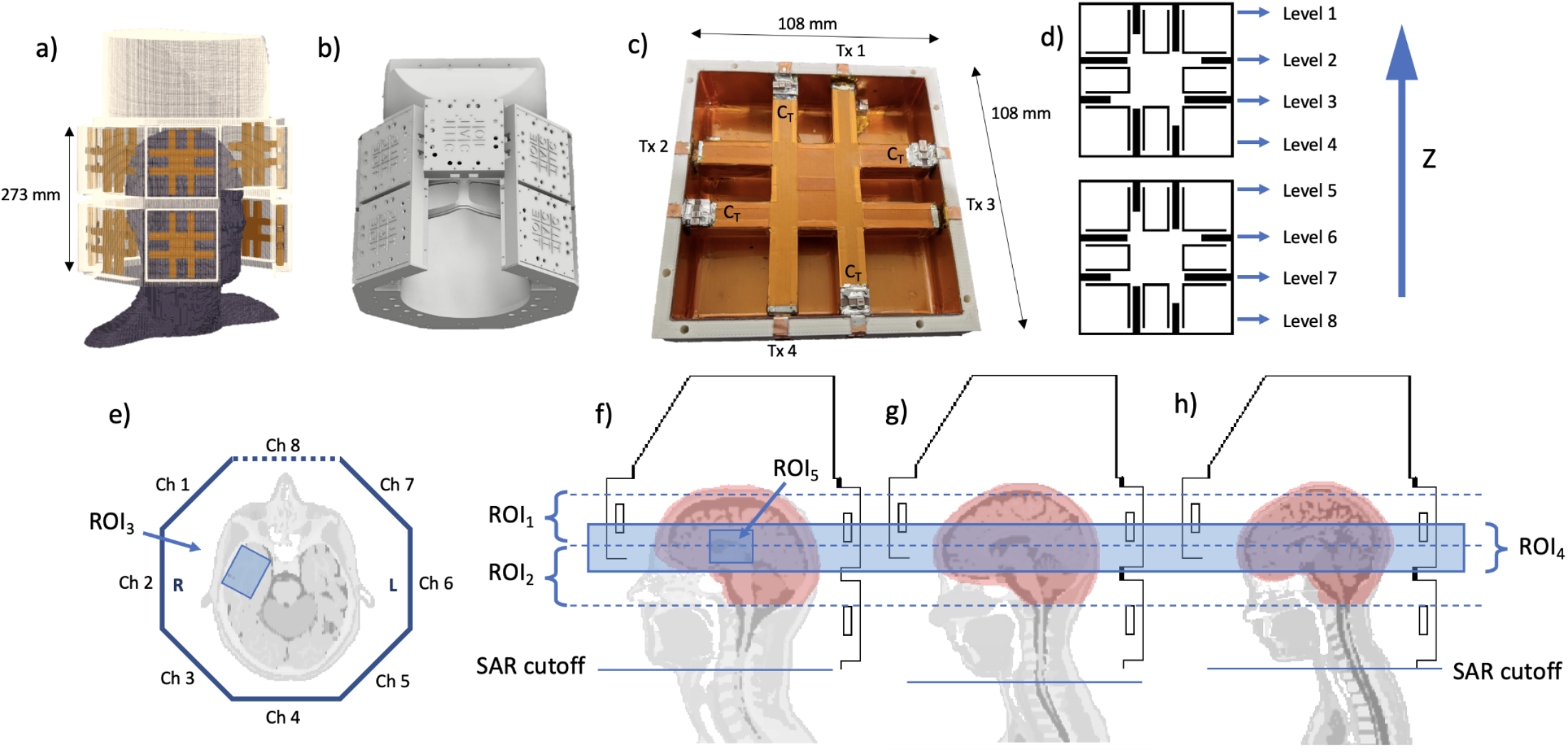
2nd generation Tic-Tac-Toe coil design: a) 3D model of the FDTD grid. b) CAD model of the coil structure. c) Single TTT panel, with mapping of transmission lines (Tx 1 – Tx 4), and tuning capacitors (C_T_). d) Cross section of the FDTD grid through the center of the panel, showing the rod positionings and the mapping of the Z-levels of the coil, where level 1 is highest in the Z direction and at the top of the head. e) Mapping of channels within each level around the coil. The top row of the coil has eight channels per level while the bottom row has seven, since it omits the panel in front of the face. f-h) Cross-section of FDTD grid showing the positions of f) Duke, g) Ella, and h) Billie in the transmit coil. Dotted lines indicate the cutoffs for ROIs 1 and 2 used in RF shimming. The blue overlays in e and f-h represent the additional ROIs (3, 4, and 5) used in the optimization of RF shim Case 3. The mask used in RF shimming and to calculate B_1_^+^ statistics is shown in f-h overlaid in red on each model. The lower bound for SAR calculations is displayed as a solid line below the chin for each head model.

### RF Simulation

The transmit coil geometry was generated using in-house code written in Python3^28^. The RF fields (B_1_^+^ and electric fields) produced by each transmit channel were simulated individually using in-house developed finite difference time domain (FDTD) software with dedicated transmission line and capacitor models^20,29^. Dimensions were 256×256×340 Yee cells, including 32 perfect matching layers on the bottom of the model (-Z), 8 at the top (+Z), and 24 on all sides (X and Y). The Yee cell size was 1.59 mm isotropic. Time resolution was approximately 3 picoseconds, and 100,000-time steps were used to reach steady state. These methods have been previously validated to provide an accurate estimation of the B_1_^+^ and electric fields^22,23,25,30^.

Anatomically detailed head models (Duke, Ella, and Billie, IT’IS Foundation Virtual Family^31^), cropped at the shoulders, were used in the simulations. The Duke model represents an adult male, age 34, weighing 70.2 kg, Ella represents an adult female, age 26, weighing 57.3 kg, and Billie represents a child female, age 11, weighing 34 kg. To compare brain size, intracranial volume for each of the models is as follows: Duke, 1.61 L; Ella, 1.57 L; Billie, 1.41 L. The models were placed within the coil considering the shoulders and as far back as possible towards the top of the receive insert. Their positions within the Tx coil are displayed in Figure 1f-h.

### Optimization

Non-subject-specific RF shimming was performed using MATLAB (MathWorks, Natick, MA) optimization functions (fmincon, GlobalSearch, interior point) to modify the amplitudes and phases of the input pulse on each channel, based on methods previously described^23^. The resolution of the B_1_^+^ fields was reduced to 3.18mm isotropic to increase the speed of the computation. For each head model, the ROI used was the whole head from the bottom of the cerebellum to the top of the brain, excluding the nasal cavities and ears, as shown as an overlay in Figure 1f-h. Coefficient of variation (CV = standard deviation/mean) of B_1_^+^ was used as the initial cost function for preliminary optimizations on the Duke model starting from random amplitudes and phases. From these cases, a few were picked with low CV and used as a starting point for further optimizations using multiple head models. The mean B_1_^+^ field in the combined ROIs and head models was constrained to produce a flip angle of at least 180° per 548V (achievable with our standard 8kW power amplifier) for 1ms square pulse to ensure adequate B_1_^+^ fields across the entire brain.

For the first RF shim case (Case 1), in order to achieve a simple, straightforward implementation, amplitudes were restricted to implementable values such that each z-level of the coil (Figure 1d) would be driven through the same splitter, i.e. the channels belonging to a z-level would have the same amplitude^23^. The power splitter configuration used in this case is outlined in Figure 2a. Phase-only RF shimming was performed using a cost function of

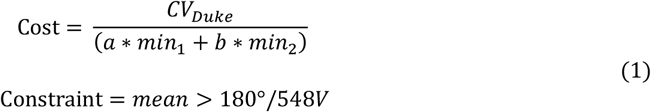

where *CV*_*Duke*_ is the coefficient of variation (CV) of B_1_^+^ in the Duke ROI, and *min*_*1*_ and *min*_*2*_ are the B_1_^+^ minimums over all three head models (Duke, Ella, and Billie) in the top half of the ROI (between the top dotted lines in Figure 1g-h, labeled as ROI_1_) and bottom half of the ROI (between the bottom dotted lines in Figure 1g-h, labeled as ROI_2_), respectively. Constants *a* and *b* are used as weights. A case was chosen with sufficient B_1_^+^ coverage by considering CV and maximum/minimum B_1_^+^ and performing visual inspection for symmetry and reduced dropout.

**Figure 2:**
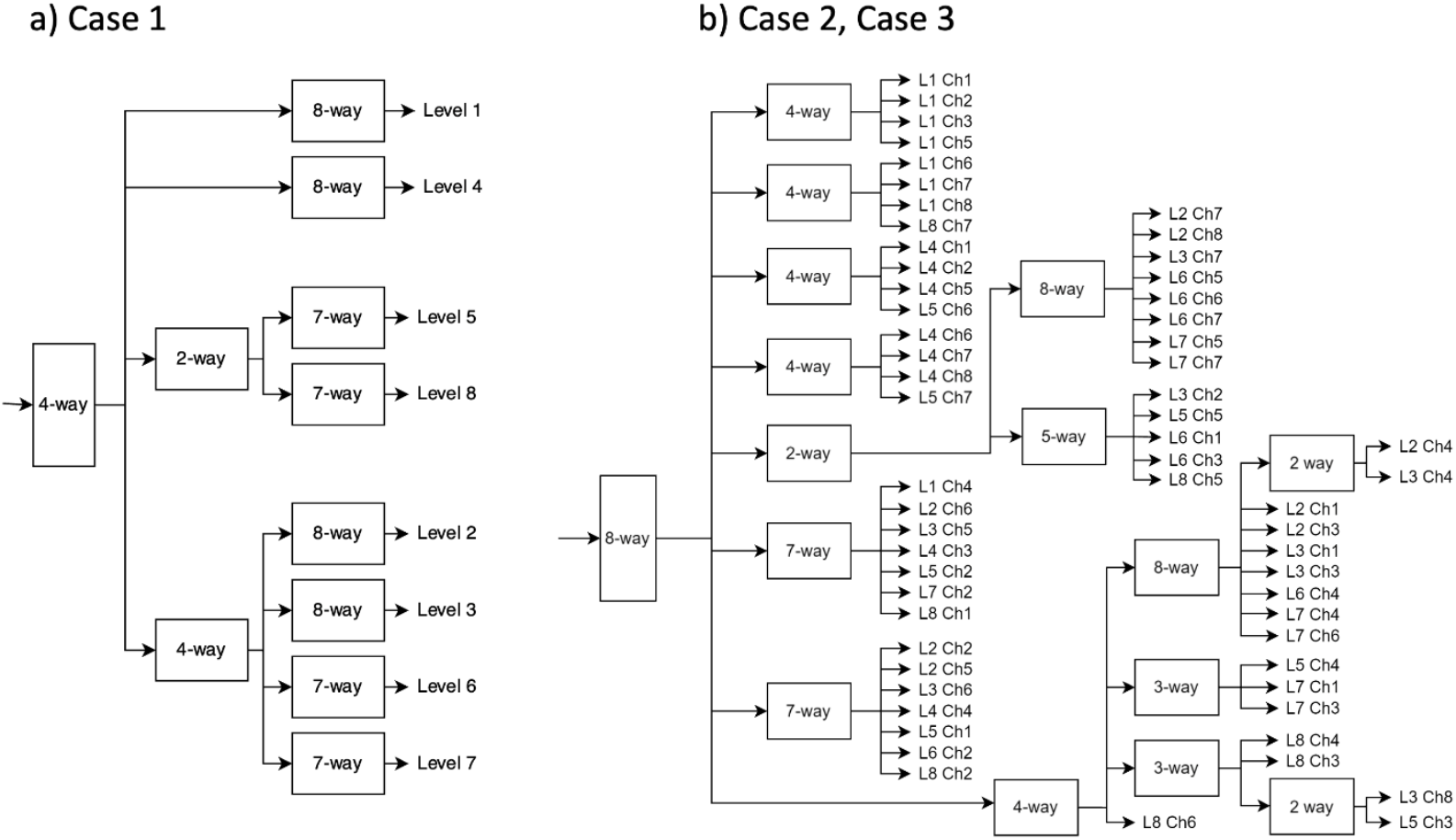
Splitter configuration for the optimized shim cases. a) Splitter configuration for RF shim Case 1, where all seven or eight channels within a z-level are held at the same amplitude and phase-only shimming is performed. b) Splitter configuration for RF shim Cases 2 and 3, which use amplitude and phase shimming. Level and channel mappings can be found in Figure 1d, e.

To generate a second, more flexible RF shim case (Case 2), a new optimization was performed, this time allowing the algorithm to modify both the amplitudes and phases on each channel. The same cost function was used (Eq. 1). A case was chosen to further extend coverage into the bottom half of the head while still providing sufficient coverage throughout. The amplitudes were rounded to the nearest implementable values; the splitter configuration used in this case is outlined in Figure 2b.

A third RF shim case (Case 3) was optimized using the same splitter configuration as in Case 2 and performing phase-only shimming, but utilizing a multi-ROI approach, isolating regions where B_1_^+^ could be improved. In addition to the same top and bottom halves of the head models used in Cases 1 and 2, two additional ROIs were placed: a slab around the axial center of the brain on all 3 models, and a small ROI around the left temporal lobe on the Duke model. These ROIs are displayed in Figure 1e-h and labeled as ROI_3_ and ROI_4_. Additionally, a small ROI was placed around the center of the brain in the Duke model (ROI_5_, Figure 1f), where the maximum B_1_^+^ typically occurs; the mean in this ROI was constrained below 270° per 548V to help keep the maximum values lower. The cost function used was

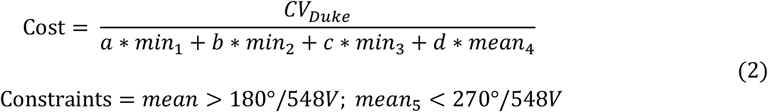

where *CV*_*Duke*_, *min*_*1*_ and *min*_*2*_ are as described above in Eq. 1, *min*_*3*_ is the minimum B_1_^+^ in the left temporal lobe of the Duke model, *mean*_*4*_ is the mean B_1_^+^ in the central slab, and *a, b, c*, and *d* are constants used for weighting.

### SAR Calculation

After optimization, SAR is calculated in MATLAB using the full resolution electric fields generated by FDTD and averaged per 10 g of tissue using the algorithm described by Caputa et al^32^. SAR statistics are calculated over the whole head, from below the chin, as designated by the solid blue lines in Figure 1f-h. This ROI is chosen to give a conservative – worst case scenario - estimate of the average SAR.

### Implementation

The coil was constructed using a 3D printed housing (ABS and polycarbonate plastic) with 17.5 μm-thick copper shielding. 6.35×6.35 mm^2^ copper rods were inserted into each leg of the TTT structure on each panel. Each channel was tuned to 297.2 MHz and matched to 50Ω using these tuning rods as well as fixed capacitors (Figure 1c). A 32-channel receive insert was 3D printed to fit within the transmit coil^33^. Using Wilkinson power splitters based on Yan et al.^34^, each of the chosen optimal RF shim cases (Figure 2) was implemented so that the coil can be used in single-transmit (sTx) mode on a Siemens 7T scanner. Coaxial cables were used to shift the phase according to the optimization output.

### Images Acquired

B_1_^+^ maps were acquired on a MAGNETOM 7T scanner (Siemens, Erlangen, Germany) in sTx mode using a Turbo-FLASH sequence^35^ with the following parameters: TR/TE = 2000/1.16ms; TA = 12 min; flip angle from 0° to 90° in 18° increments; 3.2mm isotropic resolution. The signal is fitted to a cosine function to calculate the estimated B_1_^+^ maps. B_1_^+^ maps were acquired on four healthy volunteers with informed consent as part of an approved study by the local Institutional Review Board. All procedures complied with relevant guidelines and regulations for investigational use of the device in humans.

T2-FLAIR (Fluid Attenuated Inversion Recovery) sequences were acquired on Volunteer 1 using RF shim Cases 1 and 2, with the following parameters: TE/TI/TR = 99/2900/14000 ms, resolution 0.85 × 0.85 × 1.7 mm^3^, acceleration factor 2, 5 interleaved acquisitions (64 transversal slices), TA = 8:12 min. T2-FLAIR sequences were acquired on Volunteer 2 using RF shim Cases 2 and 3, with the following parameters: TE/TI/TR = 99/2900/14000 ms, resolution 0.75 × 0.75 × 1.5 mm^3^, acceleration factor 2, 7 interleaved acquisitions for Case 2 (80 transversal slices), 8 for Case 3 (100 transversal slices), TA = 11:28 min for Case 2, 13:06 min for Case 3.

## Results

Statistical values of the simulated B_1_^+^ field and SAR are listed in Table 1 for the three RF shim cases. Case 1 provides the highest mean B_1_^+^ field in the brain mask for all three head models; Case 3 provides the lowest coefficient of variation and maximum/minimum B_1_^+^ field in the brain mask for each model. Case 1 has the highest average SAR efficiency and Case 2 has the highest peak SAR efficiency, while Case 3 has the lowest average SAR for 1W input power. Losses are included in the simulation results based on bench measurements of cables, splitters, and the plug and estimated loss in the transmit and receive coil and their components, including the TTT panels, ports, decoupling circuits, and losses in the dielectric and copper materials. Total losses ranged between 30-35% in all cases.

**Table 1:** Statistics of simulated B_1_^+^ field and SAR for three RF shim cases on three anatomically detailed head models. B_1_^+^ field statistics are calculated over each brain mask. The losses for the 2^nd^ column include losses from each case’s splitter configuration (plug, cables, boards) and from the transmit and receive coils and their components. 548 V is the maximum output for our 7T Siemens MAGNETOM (8kW power amplifier). SAR statistics are calculated over the whole head above the chin.

| | Mean $B_1^+$<br>(uT) for<br>1W input | Mean Flip Angle (°/548V) for<br>1 ms pulse width including<br>RF losses after the coil socket | CV $B_1^+$ | Max/Min<br>$B_1^+$ | Average SAR<br>Efficiency<br>($B_1^+/\sqrt{\text{SAR}_{\text{avg}}}$) | Peak SAR<br>Efficiency<br>($B_1^+/\sqrt{\text{SAR}_{\text{peak}}}$) | Average SAR<br>(W/kg) for<br>1W input | Peak SAR<br>(W/kg) for<br>1W input | Peak/Avg<br>SAR |
| --- | --- | --- | --- | --- | --- | --- | --- | --- | --- |
| **Case 1** |  |  |  |  |  |  |  |  |  |
| **Duke** | 0.348 | <u>209.57</u> | .231 | 4.42 | <u>1.55</u> | 0.867 | 0.051 | 0.161 | 3.19 |
| **Ella** | 0.330 | 198.32 | .217 | 4.32 | 1.43 | 0.867 | 0.053 | 0.144 | 2.74 |
| **Billie** | 0.344 | 207.02 | .219 | 3.51 | 1.40 | 0.860 | 0.060 | 0.160 | 2.66 |
| **Case 2** |  |  |  |  |  |  |  |  |  |
| **Duke** | <u>0.373</u> | 203.64 | .210 | 4.58 | 1.52 | 0.857 | 0.060 | 0.189 | 3.17 |
| **Ella** | 0.349 | 190.40 | .216 | 4.43 | 1.40 | <u>0.873</u> | 0.062 | 0.159 | <u>2.59</u> |
| **Billie** | 0.358 | 195.54 | .220 | 3.58 | 1.36 | 0.776 | 0.069 | 0.213 | 3.08 |
| **Case 3** |  |  |  |  |  |  |  |  |  |
| **Duke** | 0.324 | 184.55 | <u>0.197</u> | 4.06 | 1.54 | 0.790 | <u>0.044</u> | 0.169 | 3.80 |
| **Ella** | 0.308 | 175.61 | 0.198 | 3.92 | 1.42 | 0.825 | 0.047 | <u>0.140</u> | 2.97 |
| **Billie** | 0.317 | 180.75 | 0.204 | <u>3.49</u> | 1.37 | 0.774 | 0.054 | 0.171 | 3.15 |

Figure 3 shows the simulated B_1_^+^ maps on each of the three head models for each of the three finalized RF shim cases. Arrows highlight areas of improvement across the three shim cases, particularly in the cerebellum and left temporal lobe, and histograms are plotted of the B_1_^+^ field in each of the head masks. From Case 1 to Case 3, histograms become narrower with fewer low values. SAR maps for each case and each head model are displayed in Figure 4, shown in W/kg for 1W input power, averaged over 10g.

**Figure 3:**
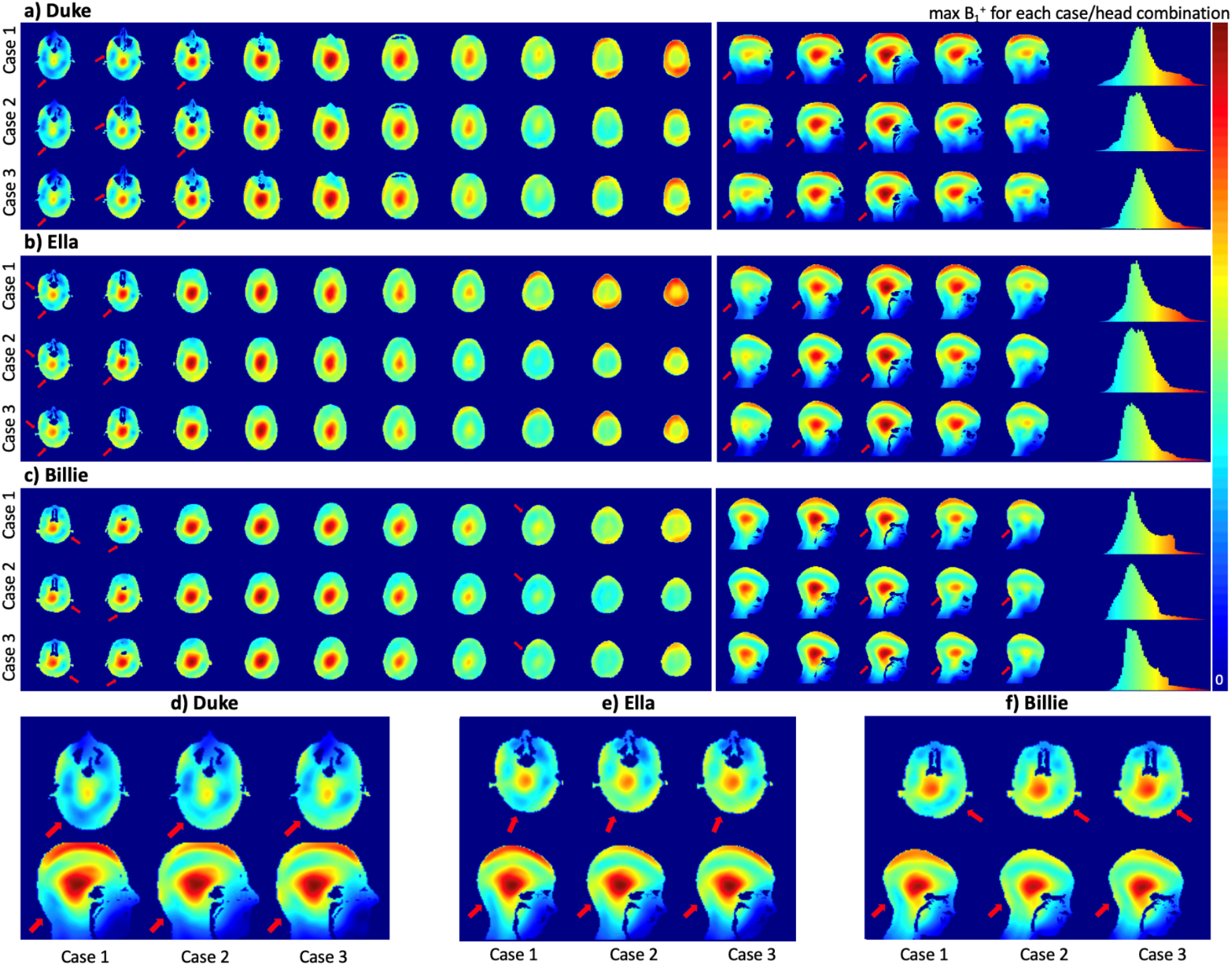
Simulated B_1_^+^ maps in three head models using three RF shim cases: a) Duke model, Cases 1, 2, and 3 from top row down; b) Ella model, Cases 1, 2, and 3; c) Billie model, Cases 1, 2, and 3. For each model/case, the B_1_^+^ map is scaled to from zero to the maximum B_1_^+^ for that model/case. Histograms demonstrate the B_1_^+^ field distribution for each model/case within the brain mask, from the top of the head to bottom of cerebellum. Zoomed-in views of the bottom axial slice (through the cerebellum) and center sagittal slice are shown for each head model; d) Duke, e) Ella, and f) Billie; and for shim Cases 1, 2, and 3 from left to right.

**Figure 4:**
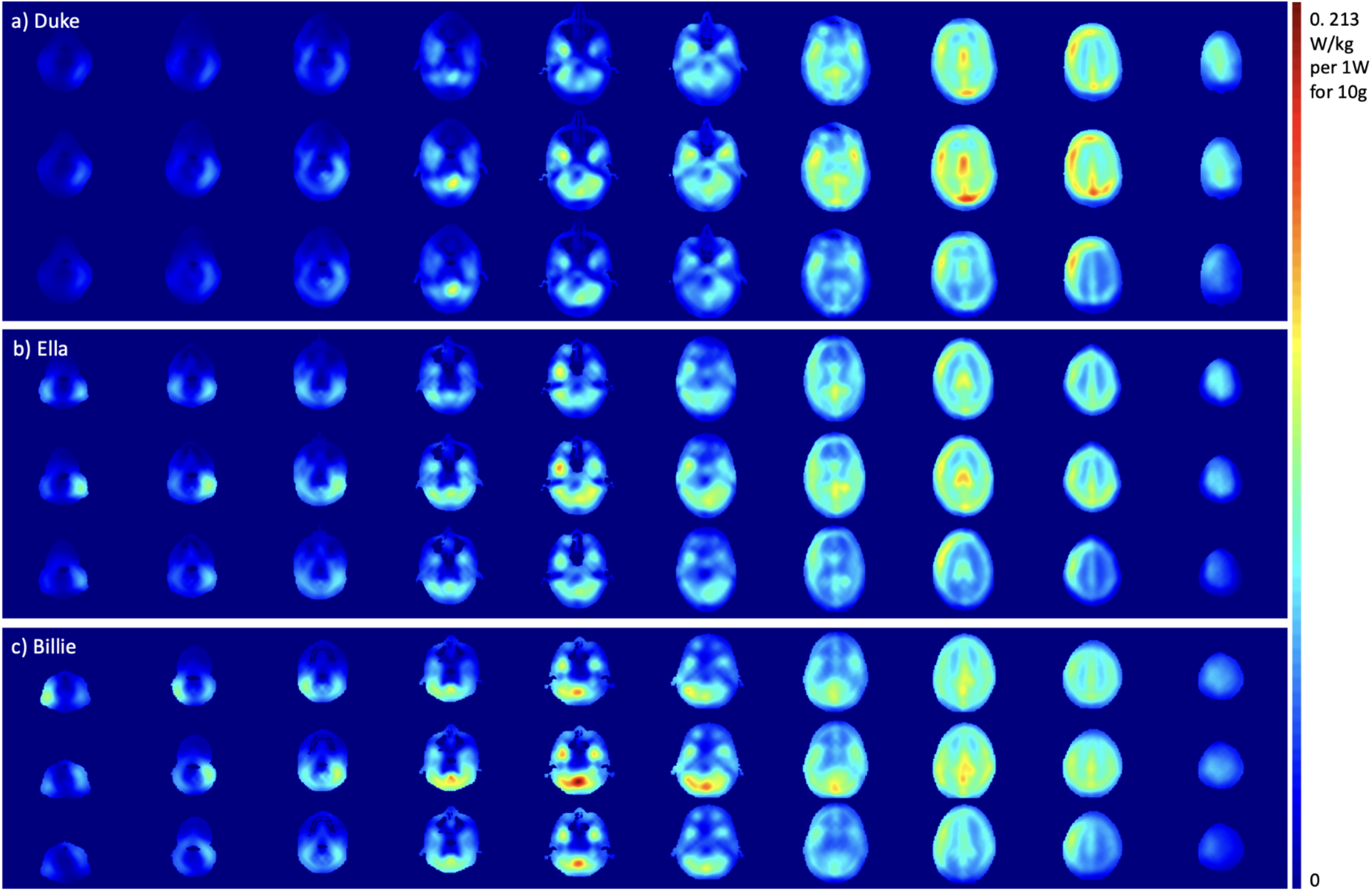
Simulated SAR per 1W in three head models using three RF shim cases, averaged over 10g of tissue: a) Duke model, Cases 1, 2, and 3 from top row down; b) Ella model, Cases 1, 2, and 3; c) Billie model, Cases 1, 2, and 3. All cases/models are scaled to the same maximum value of 0.213 W/kg per 10g of tissue (observed on the Billie model for Case 2) for 1W input power.

Figure 5 shows the B_1_^+^ field distribution in vivo in Volunteer 1, who was scanned with all three RF shimming cases, as well as three volunteers (2-4) who were scanned with RF shim Case 2 and Case 3. All volunteers were between 25-29 years old; one subject was male and three were female. Volunteer intracranial volumes are as follows: Volunteer 1, 1.46 L; Volunteer 2, 1.67 L; Volunteer 3, 1.57 L; Volunteer 4, 1.47 L. Figure 5 also shows histograms of B_1_^+^ values in the brain mask, and mean and CV values are also displayed for each volunteer. Mean B_1_^+^ decreases from Cases 1 and 2 to Case 3 but fewer lower values (typically observed in cerebellum and/or temporal lobes) are achieved where the CV also improves. Figure 6 shows FLAIR images (raw from the scanner) on Volunteer 1 (using Cases 1 and 2) and Volunteer 2 (using Cases 2 and 3). Arrows show regions of improvement between the cases.

**Figure 5:**
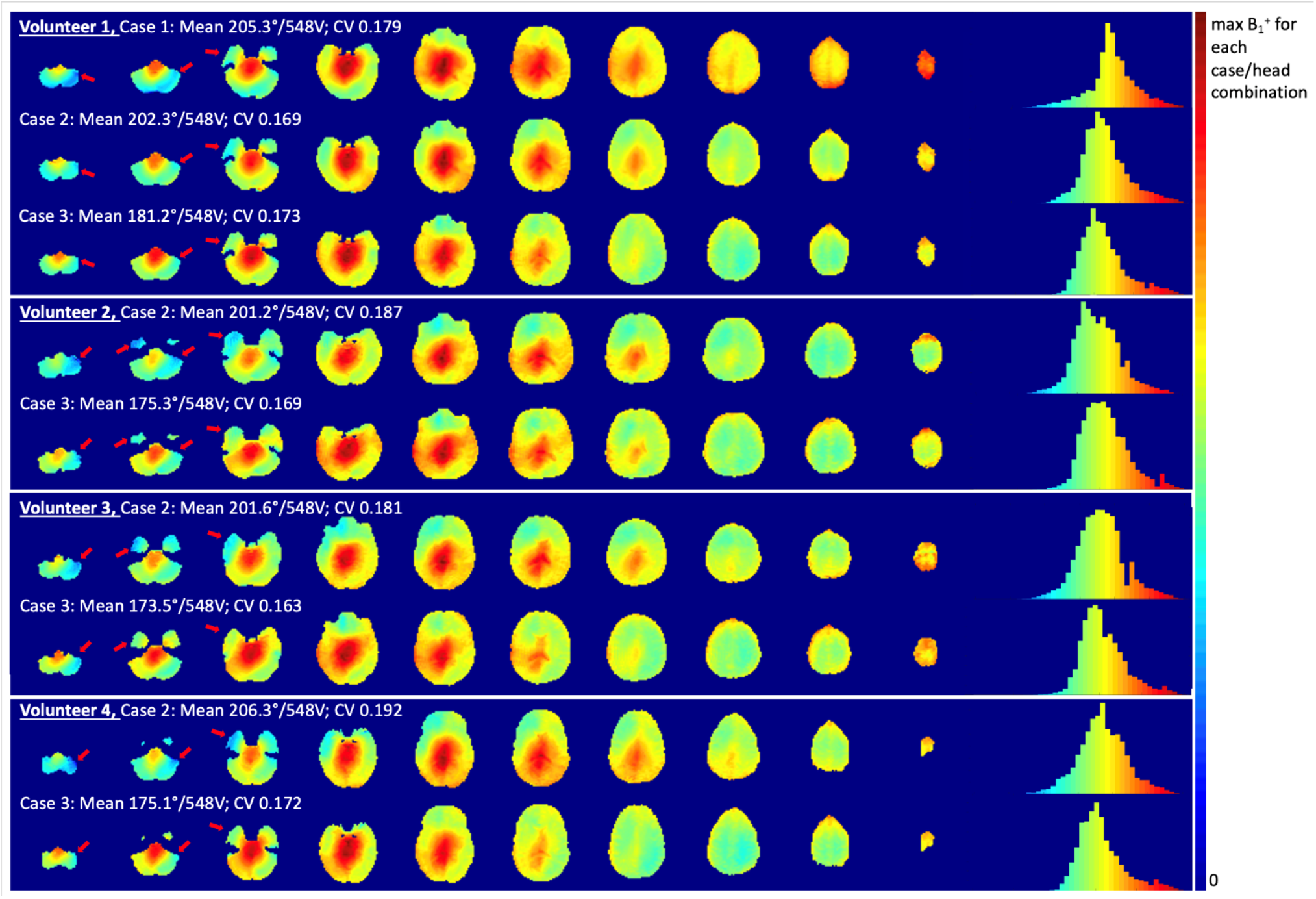
In vivo B_1_^+^ maps on volunteers who were scanned using the RF shim cases: one volunteer who was scanned using all three RF shim cases, and three volunteers using only Case 2 and Case 3. Each case/volunteer is scaled to its own maximum B_1_^1+^. Histograms demonstrate the B_1_^+^ distribution across the brain for each volunteer/shim case, with the x-axis going from zero to the maximum B_1_^+^ for that head model/case combination. The flip angle values assume 1 ms pulse width.

**Figure 6:**
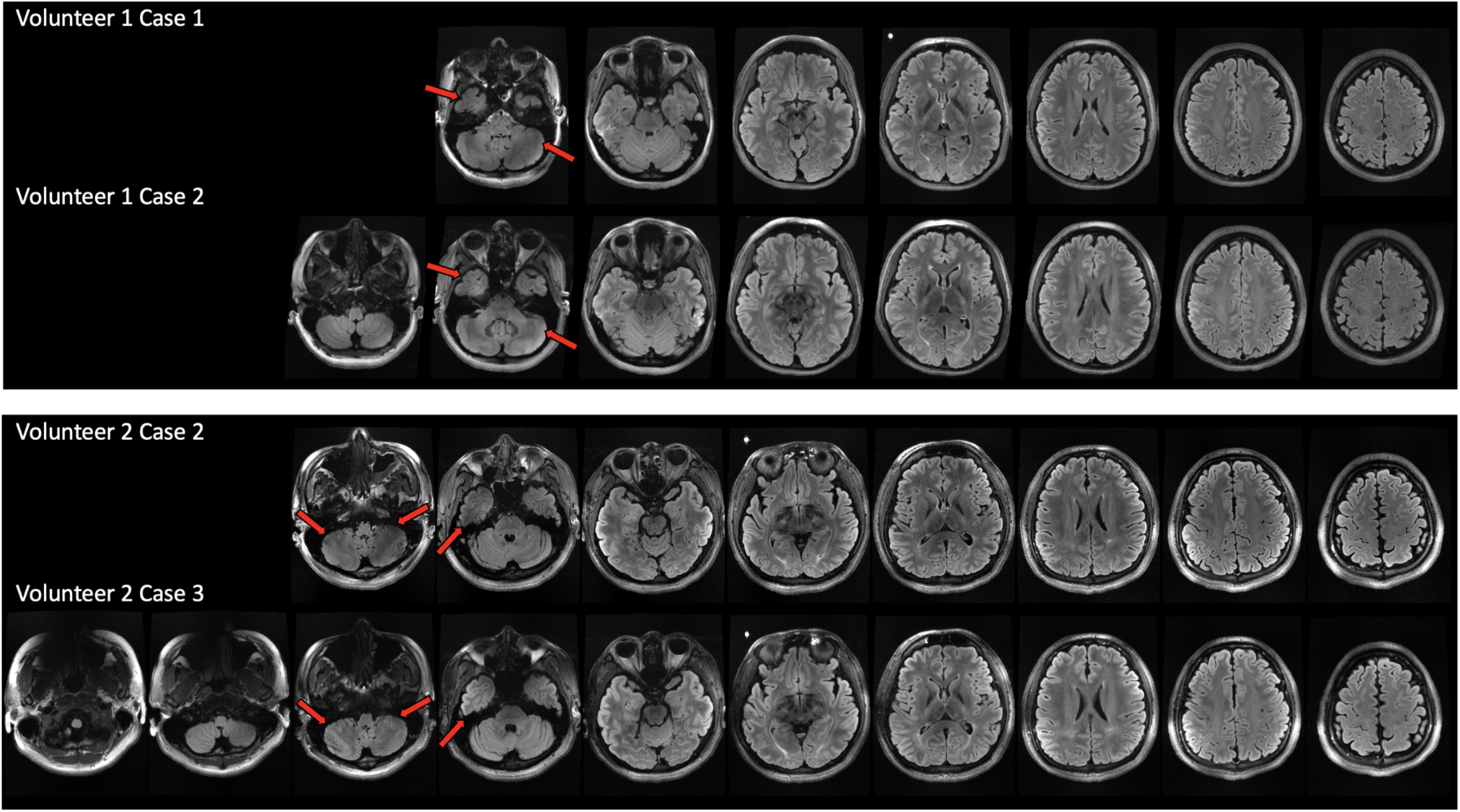
T2-FLAIR images (raw from the scanner) acquired on volunteers 1 and 2 using the 60-channel Tx/32-channel Rx TTT RF coil. The sequence used for Volunteer 1 was 64 slices, 0.85 × 0.85 × 1.7 mm resolution. The sequence used for Volunteer 2 was 0.75 × 0.75 × 1.5 mm resolution; Case 2 was 80 slices and Case 3 was 100 slices. Volunteer 1 has an ICV of 1.46 L, head dimensions of 19 cm AP, 16.2 cm RL, and 14.7 cm from the top of the skull to the bottom of the cerebellum. Volunteer 2 has an ICV of 1.67 L, head dimensions of 19.5 cm AP, 17.7 cm RL, and 15.2 cm from the top of the skull to the bottom of the cerebellum.

## Discussion

In this work, we present RF shimming techniques for optimizing the 2^nd^ Generation Tic-Tac-Toe RF Coil System for routine use with the sTx systems at 7T. The increased number of transmit channels (60) improves efficiency in the azimuthal direction and provides more degrees of freedom (2*n* -1 for *n* channels) for RF shimming as compared to the 1^st^ Generation TTT RF Coil System with fewer transmit channels (16). To develop a robust and homogeneous implementation on the sTx system, three head models were used simultaneously, chosen to better reflect the demographics of the subjects scanned in ongoing studies at our facility. Optimizing over three head models of different sizes and positions enables the coil to perform well for a range of ages and head types while utilizing the sTx mode of the scanner.

In addition to utilizing three head models in RF shimming, a multi-ROI strategy was used. The first two shim cases developed in this work utilized a dual-ROI strategy which aimed to improve B_1_^+^ field homogeneity in the lower regions of the brain (temporal lobe, brainstem, cerebellum) which are often difficult to excite at 7T regardless of coil design while maintaining low CV across the head. The third shim case used several additional ROIs to isolate regions that should be further improved, increasing signal in the temporal lobe and improving homogeneity across the central parts of the brain. The ROIs used may be specific to our coil, but these are regions that are often problematic in other coil designs at 7T due to the wavelength and dielectric artefacts. Therefore, the same strategy can be applied to the optimization of any RF transmit array with small adjustments to the locations of the ROIs. Figure 3 highlights the improvements in simulation between shim Cases 2 and 3, demonstrating the benefits of a multi-ROI shimming strategy. These improvements are reflected in the in-vivo field maps and images shown in Figures 5 and 6. This shimming framework is flexible to changing or adding ROIs and/or terms in the cost function, allowing for adjustments with coil geometry changes, and regions of too-low or too-high B_1_^+^ field can be specifically targeted and improved.

Furthermore, the power splitting strategy can largely impact B_1_^+^ and SAR. Case 1 and Case 2 used the same shimming method (i.e. the same cost function and ROIs) while using different power splitter configurations. As Case 1 was the earliest implementation of the coil, we fixed the amplitudes according to z-level, assigning more power to levels which had larger impact on overall B_1_^1+^field. For Case 2, by allowing free amplitude-and-phase shimming, even using the same cost function, the B_1_^+^ field distribution significantly improved. Case 3 uses the multi-ROI cost function but uses phase-only shimming so that we could reuse the splitters implemented in Case 2. Further improvements could be further achieved using a multi-ROI shimming strategy and allowing for phases and amplitudes to vary.

The RF shimming cases developed in this work were implementable on the coil by using power splitters and coaxial cables of different lengths. The splitter configurations are shown in Figure 2; Case 1 utilizes a total of 11 power splitters, while the configuration used by both Cases 2 and 3 requires 16 splitters. In vivo B_1_^+^ maps demonstrate the improvements in homogeneity and extended coverage seen with Cases 2 and 3. Since Case 1 was only in use for a limited amount of time, only one volunteer had been scanned using all three cases. Three volunteers were scanned with Case 2 and Case 3. FLAIR images similarly show the improvements in B_1_^+^ field across the three cases. Using Case 3 allows us to achieve improved coverage below the cerebellum.

In order to achieve improved B_1_^+^ field homogeneity in Case 3, some tradeoffs must be made in overall efficiency. Nonetheless, the coil is still able to provide sufficient B_1_^+^ field intensity with an average flip angle from simulations and experiments of ∼ 180° with 1ms pulse width at the scanner’s allowed voltage. Additionally, the ratio of peak to average SAR is highest in Case 3, demonstrating another tradeoff in order to achieve homogeneous B_1_^1+^field, although the SAR levels are still relatively low^25^. In this work, we did not include a SAR parameter in our optimization cost function. The addition of a SAR parameter in the cost function could allow the optimization software to balance homogeneity and SAR^23,36^. Even without the inclusion of the SAR in the cost function, safe levels of SAR are achieved across all models and in all three cases presented here.

## Conclusion

The presented RF shimming techniques implemented on the 2^nd^ Generation TTT RF Coil System demonstrate the ability to provide non-subject-specific homogeneous B_1_^+^ field excitation for routine neuroimaging at 7T MRI on the sTx mode. The 60-channel design improves power efficiency and enables a higher degree of RF shimming flexibility, achieving extended brain coverage while maintaining patient comfort with an open visual field. Three optimized non-subject specific RF shim cases, developed using multiple head models, exhibit improvements in homogeneity and coverage, particularly in challenging-to-image regions such as the temporal lobes and cerebellum at 7T. These RF shim cases also show safe SAR levels across all models, making this coil system an effective tool for routine 7T neuroimaging, already with > 1750 in-vivo human scanning sessions over the past 28 months.

## Data Availability

All data produced in the present study are available upon reasonable request to the authors.

## Acknowledgement

This work was supported by the National Institutes of Health under award numbers R01MH111265, R01AG063525, R56AG074467, and T32MH119168, and by the National Science Foundation Graduate Research Fellowship under Grant No. 1747452. This research was supported in part by the University of Pittsburgh Center for Research Computing, RRID:SCR_022735, through the resources provided by NSF award number OAC-2117681 and NIH award number S10OD028483. We also thank Dr. Boris Keil for providing the CAD model for the receive insert, Dr. Tiago Martins for creating the code to generate the FDTD model of the transmit coil, and Bruno DeAlmeida and Tobias Campos for their assistance and support with this project.

